# ASSOCIATION BETWEEN CHANGES IN PSYCHOLOGICAL READINESS AND SUBJECTIVE KNEE FUNCTION AFTER ACL RECONSTRUCTION

**DOI:** 10.64898/2026.03.06.26347201

**Authors:** Olivia Johnson, Colin W. Bond, Benjamin C. Noonan

## Abstract

**Background:** Psychological readiness to return to sport and subjective knee function are critical outcomes following ACL reconstruction (ACLR), yet they do not always progress in parallel. An athlete may demonstrate high subjective knee function but low psychological readiness, suggesting a mental barrier to return, or conversely, report high readiness despite persistent functional limitations, raising concerns of overconfidence and reinjury risk. Understanding how these domains change together during recovery is essential for identifying mismatches that may require targeted intervention.

**Purpose:** The purpose of this study is to examine the relationship between changes in psychological readiness (ACL-RSI) and subjective knee function (IKDC) from early to late recovery following ACLR.

**Study Design:** Secondary analysis of prospectively collected data.

**Methods:** Athletes (N = 48, Age at ACLR = 17.7 ± 1.8 y) aged 15-25 years who underwent ACLR with an ipsilateral autograft, had a pre-injury MARX score > 8, and completed the ACL-RSI and IKDC questionnaires at 3.5 ± 1 and 7 ± 1 months post-ACLR were included. Percent changes in ACL-RSI and IKDC scores between early and late recovery were calculated. Spearman’s rank correlation was used to examine the association between changes in psychological readiness and subjective knee function. Significance was set to p < .05.

**Results:** The mean percent change in ACL-RSI was 40.7 ± 57.1% and the mean percent change in IKDC was 24.8 ± 18.1% from 3.5 ± 1 months to 7 ± 1 months post-ACLR. The percent changes in ACL-RSI and IKDC scores from 3.5 ± 1 months to 7 ± 1 months post-ACLR were moderately correlated (ρ = 0.350 (95% CI [0.089, 0.584]), p = 0.012).

**Discussion:** The main finding of this study was that subjective knee function and psychological readiness to return to sport changed in parallel from 3.5 to 7 months following ACLR. Clinicians can use this information regarding the concordant progression of psychological readiness to return to sport and subjective knee function to personalize ACL rehabilitation for future patients. Overall, clinicians can understand that if psychological readiness improves, subjective knee function will likely improve over the 3.5- to 7-month post-ACLR time frame, and vice versa. Therefore, focusing on both of these components at multiple time points during the recovery process may be influential to ensure the greatest likelihood of returning to sport in athletes following ACLR.

## INTRODUCTION

Patient-reported outcomes (PROs) after anterior cruciate ligament reconstruction (ACLR), such as psychological readiness to return to sport (RTS) and subjective knee function, are associated with patient prognosis, yet how these domains change in tandem over the course of rehabilitation and RTS is not well understood. Although the focus of ACLR is on physical rehabilitation, a growing body of research highlights the importance of psychological readiness, one of the most widely studied constructs following ACLR, encompassing an athlete’s confidence, emotional response, and fear of reinjury^1,4,7,11,13,15-17^. Lower psychological readiness has been linked to poorer outcomes, including delayed or unsuccessful RTS^3,6,9^. Similarly, reduced subjective knee function has been associated with lower activity levels and fear of reinjury^4,12^. From a patient’s perspective, one’s perception of knee function may influence psychological readiness^2^; however, how these domains progress in tandem over time post-ACLR remains unknown.

PROs, including the ACL-Return to Sport Index (ACL-RSI) and the International Knee Documentation Committee Subjective Knee Function scale (IKDC), measure psychological readiness to RTS and subjective knee function, respectively^1-3, 5-6, 8-9^. The ACL-RSI focuses on an athlete’s psychological readiness to resume sports participation^17-18^, specifically on performance confidence, risk appraisal, and fear of reinjury, while the IKDC evaluates subjective knee function, including pain, stability, and performance^19^.

Research shows that scores on these PROs have a strong positive correlation to each other when assessed concurrently at a single point in time, 6 months post-ACLR^16^, suggesting that an athlete’s perception of their knee function is strongly associated with their psychological readiness^2^. Furthermore, several studies have also demonstrated that scores on each PRO improve independently over time post-ACLR. For example, ACL-RSI improved over 6, 12, and 24 months after ACLR^6,9,17^ and IKDC scores also improved from 3 to 12 months following ACLR^12^. However, it remains unclear how ACL-RSI and IKDC scores progress in tandem over time post-ACLR—that is, whether changes in one are proportionally associated with changes in the other across time post-ACLR.

Ohji et al. investigated whether changes in ACL-RSI and IKDC scores were proportionally related from pre-ACLR and 6 months post-ACLR. The study found that both ACL-RSI and IKDC improved independently, but the magnitudes of improvement were not correlated^8^. One potential explanation for the lack of correlation is that psychological readiness to RTS, as measured by the ACL-RSI, may not be fully developed or meaningful when assessed pre-ACLR. However, another study using a specific secondary prevention training program found that athletes who demonstrated greater improvements in ACL-RSI also reported better IKDC scores, although the study did not directly assess whether changes in the two scores were proportionally related over time^5^. These conflicting results indicate a need for a clearer understanding of how subjective knee function, as measured by IKDC, and psychological readiness, as measured by ACL-RSI, progress in tandem over time post-ACLR. This may allow clinicians to better individualize rehabilitation strategies that target mismatches between subjective knee function and psychological readiness based on a patient’s unique recovery profile, thereby optimizing outcomes following ACLR. For example, some athletes may demonstrate a large increase in subjective knee function but smaller changes in psychological readiness, suggesting a mental barrier to return, or conversely, report large increases in psychological readiness or consistently high psychological readiness despite persistent functional limitations, raising concerns of overconfidence and reinjury risk.

Therefore, the purpose of this study is to analyze the progression of ACL-RSI and IKDC in tandem among adolescent and young adult athletes following ACLR by evaluating the proportional change in scores between 3.5 ± 1 months and 7 ± 1 months post-ACLR. We hypothesize that the change in ACL-RSI and IKDC scores between 3.5 ± 1 months and 7 ± 1 months post-ACLR will demonstrate a weak, positive correlation.

## METHODS

### Study Design and Participants

The present investigation was an observational secondary analysis of prospectively collected data. Beginning in 2012, post-ACLR return-to-sport (RTS) test data from athletes who completed RTS testing at our health system’s sports physical therapy clinics were prospectively entered into a digital clinical data repository managed using REDCap. Anonymous data were obtained from this repository by an honest broker, which contained records for 1,187 patients as of October 2025. Athletes who met all of the following criteria were included in the analysis: (1) age 15–25 at the time of injury; (2) pre-injury MARX score > 8; (3) ACLR performed using an ipsilateral autograft; and (4) completion of both the ACL-RSI and IKDC scales at 3.5 ± 1 months and 7 ± 1 months post-ACLR. Athletes who met one or more of the following criteria were excluded from the analysis: (1) multi-ligament reconstruction involving the MCL, LCL, or PCL; (2) additional procedures such as manipulation under anesthesia or lysis of adhesions following the initial ACLR; (3) history of previous ACL injury; or (4) a delay of more than 12 months between ACL injury and ACLR. The Sanford Health Institutional Review Board granted this study an exemption (Study ID: 2390).

### Data Collection Procedures

Athletes in the data repository may have undergone rehabilitation within our health system or may have been referred from outside health systems or physical therapy clinics. Some athletes completed only portions of their rehabilitation with our providers or underwent RTS testing exclusively at our clinics, whereas the majority participated in rehabilitation under our health system’s criterion-based, individualized protocol. This protocol initiated early, progressive weight-bearing immediately post-operatively in the absence of concomitant procedures, with brace use and progression determined by quadriceps control and joint stability. Rehabilitation advanced through staged phases emphasizing restoration of range of motion, neuromuscular control, strength, and movement quality. RTS tests were administered to guide progression and support RTS decision-making, typically around 3.5 and 7 months post-ACLR, with additional tests completed at the provider’s discretion. RTS tests may include a variety of physical, clinical, and PRO items.

### Subjective Knee Function

Athletes completed the IKDC as part of their RTS test. The IKDC is a 19-item instrument that evaluates knee symptoms, functional status, and limitations in sports or physical activity^19^. Each item is assigned a point value, and the total score is converted to a standardized scale ranging from 0 to 100, where higher scores reflect fewer symptoms and less dysfunction and lower scores reflect more symptoms and greater dysfunction.

### Psychological Readiness

Athletes completed the ACL-RSI as part of their RTS test. The ACL-RSI is a 12-item questionnaire that assesses psychological readiness to return to sport after ACLR^17^. The scale evaluates three domains including emotions, confidence in performance, and risk appraisal. Item scores are averaged to produce a total score ranging from 0 to 100, with higher scores indicating greater psychological readiness to RTS and lower scores representing lower psychological readiness to RTS.

### Statistical Analysis

Each athlete’s ACL-RSI and IKDC scores were used to calculate percent changes from 3.5 ± 1 months to 7 ± 1 months post-ACLR; these percent changes were the primary variables for the analysis. The association between percent change in ACL-RSI scores and percent change in IKDC scores was examined using Spearman’s rank correlation. This non-parametric method was employed to account for potential non-linear relationships and non-normal percent change distributions. Additionally, we employed a non-parametric bootstrap approach using 10,000 resamples with replacement to estimate a 95% confidence interval for Spearman’s ρ. Significance was set to p < 0.05. To address the stated research question and hypothesis, an a priori sample size calculation indicated that a minimum of 194 athletes would be required to detect a weak, positive correlation (ρ = 0.20) with 80% power at a two-sided significance level of 0.05.

## RESULTS

After exclusions, 27 male and 21 female patients (N = 48, age at ACLR = 17.7 ± 1.8 y) remained available for analysis. The mean percent change in ACL-RSI was 40.7 ± 57.1% and the mean percent change in IKDC was 24.8 ± 18.1% from 3.5 ± 1 months to 7 ± 1 months post-ACLR. The Spearman rank correlation revealed that the percent changes in ACL-RSI and IKDC scores from 3.5 ± 1 months to 7 ± 1 months post-ACLR were moderately correlated (ρ = 0.350 (95% CI [0.089, 0.584]), p = 0.012) (Figure 1).

**Figure 1.**
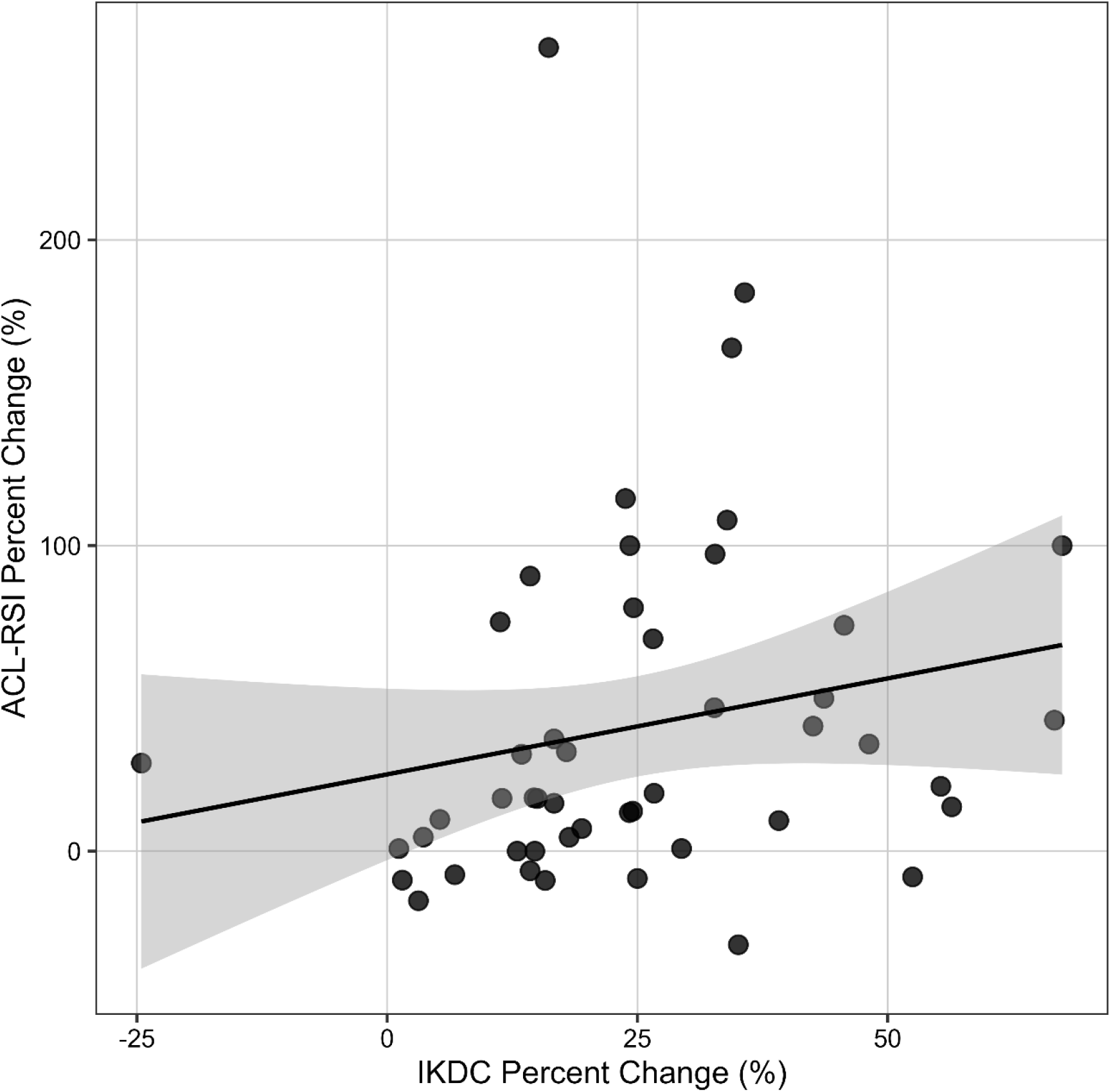
The association between the Anterior Cruciate Ligament–Return to Sport Index (ACL-RSI) percent change and the International Knee Documentation Committee Subjective Knee Function scale (IKDC) percent change from 3.5 ± 1 months to 7 ± 1 months post-anterior cruciate ligament reconstruction. Black line: linear regression; grey band: 95% confidence interval for the linear regression; black dots: individual patients.

## DISCUSSION

Psychological readiness, as measured by ACL-RSI, and subjective knee function, as measured by IKDC, are key PROs that may help personalize treatment plans for athletes recovering from ACLR. The hypothesis of this study was that the change in ACL-RSI and IKDC scores between 3.5 ± 1 months and 7 ± 1 months post-ACLR would demonstrate a weak, positive correlation. The main finding of this study was that subjective knee function and psychological readiness to return to sport changed in parallel from 3.5 to 7 months following ACLR. Therefore, these findings indicate that changes in psychological readiness and subjective knee function are concordant, supporting a meaningful association during this stage of recovery.

The results of our study indicate that there was a smaller mean percent change in IKDC than the mean percent change in ACL-RSI from 3.5 months to 7 months post-ACLR. Likewise, all IKDC percent changes were positive, indicating that every athlete demonstrated improvement in subjective knee function from 3.5 to 7 months post-ACLR. Notably, 25% of the athletes had either ACL-RSI scores that remained the same or decreased from 3.5 to 7 months post-ACLR. This may suggest that it takes a longer recovery time to improve psychological readiness to RTS than subjective knee function in athletes. A possible explanation for this pattern is that an athlete’s perception of knee strength and pain may outweigh their belief that they are ready to return to prior sport participation. Additionally, another explanation may be that patients were analyzed before RTS and before incorporating sport-specific movements into rehabilitation. These athletes may have the subjective idea that their knee is not painful and stable; however, without incorporating similar movements in rehabilitation to their sport at 7 months, they may not feel psychologically ready to return to sport. Although they are positively correlated when measured concurrently, the timing of improvement between the two constructs may differ, which could be valuable to investigate in future studies.

Interestingly, these results disagree with the previously mentioned study by Ohji et al., in which ACL-RSI and IKDC scores independently improved but had no association with each other from pre-ACLR to 6 months post-ACLR^8^. In comparison to our findings following ACLR, this may indicate that measuring ACL-RSI and IKDC before ACLR may not have predictive value for psychological readiness to RTS. A possible explanation for these differences could be due to various factors. As one example, before surgery an individual may be psychologically ready to return to sport, possibly from being passionate about their sport or not coming to terms with their injury yet, but may perceive their knee as being unstable and painful, due to the knowledge that they are undergoing ACLR. Ohji et al. found that those with low ACL-RSI before ACLR didn’t have significant improvement after 6 months post-ACLR; however, there were no significant changes among those who started with high ACL-RSI results^8^. It is possible that ACL-RSI, administered pre-ACLR, is a proxy for resilience^20^. For example, an athlete could be acknowledging their return to sport at some point in the future, or their resilience, rather than their readiness to RTS at that exact moment. Specifically, athletes who started with high ACL-RSI results pre-operatively could have higher resilience, but don’t demonstrate an increase in ACL-RSI shortly after ACLR. Additionally, differences between male and female athletes in relation to psychological readiness could have an impact on the changes in study results. The majority of athletes in the Ohji study were female athletes^8^. This could provide an explainable difference if male athletes had higher ACL-RSI scores in the initial stages of rehabilitation before converging with female athletes’ scores at approximately 5 months post-ACLR, which our lab has previously studied^21^. Future research could continue to evaluate whether each sex has distinct responses in ACL-RSI and IKDC scores, addressing this uncertain variable. Overall, these results indicate that starting psychological rehabilitation post-ACLR, focusing on subjective knee function and psychological readiness to RTS, is essential and could play a large role in an athlete’s recovery following ACLR.

Although the findings were dissimilar to the Ohji et al. study, another study found a positive association between ACL-RSI and IKDC while using the ACL-SPORTS Training Program at 1 and 2 years following ACLR^5^. As shown in Figure 1, the majority of athletes in this study demonstrated a significant association between ACL-RSI and IKDC scores, and the changes between the two were positively correlated. The data from this study were collected at an earlier recovery period than in the ACL-SPORTS training program. However, the data from both studies may suggest that psychological recovery may require a prolonged period regardless of a specific training program.

It is notable to mention that the ACL-RSI and IKDC questionnaires likely do not change in isolation. As patients get stronger, move better, and tolerate higher level rehabilitation drills, they often report less pain and more stability, and they also tend to feel more confident about RTS. In other words, improvements in ACL-RSI and IKDC may rise alongside other objective recovery markers such as quadriceps strength, hop performance, swelling, and activity exposure. This may help clinicians see PROs as part of a larger recovery picture, not as stand-alone scores. Additionally, this dataset likely includes some patients who later tore their ACL again, either in the same knee or the opposite knee. If those cases are present, future work could compare patients with injuries post-ACLR to those without and ask a practical clinical question: do patients who show a mismatch between IKDC and ACL-RSI have a higher reinjury risk? For example, a patient who feels very ready but still reports poor knee function might be holding back and not preparing for sport demands. Looking at the two PROs together, rather than interpreting each one alone, could help flag patients who need targeted physical training, targeted psychological support, or both before returning to sport.

It is apparent that testing psychological readiness and subjective knee function at multiple time points throughout recovery following ACLR is important. For instance, if these two variables are only measured at one time point, the clinician may never know if rehabilitation is progressing as expected, specifically that ACL-RSI and IKDC are progressing in the same direction. Scores at one time point may be marginally important, but how scores progress together over the course of rehabilitation may be more crucial. Moreover, it is likely that athletes will present to rehabilitation after ACLR with various levels of psychological readiness and subjective knee function, clearly an important aspect of recovery that all providers should personalize. For example, if there is a large improvement in IKDC but minimal improvement in ACL-RSI in an athlete, the individual may believe their knee is healthy and functioning well but still not be able to RTS. Conversely, if an athlete has a large improvement in psychological readiness but minimal improvement in IKDC, they may RTS with a sub-optimal functioning knee. Although most of the athletes in our study had some level of improvement in IKDC, it is still likely that athletes across the country may present with lower IKDC levels and may need additional physical rehabilitation before RTS. The results of this study, specifically that ACL-RSI and IKDC improve together, indicate that using at least one of the PROs to implement during the rehabilitation period could be a valuable strategy for clinicians. Specifically, implementing multiple aspects of psychological readiness in ACLR recovery may be more beneficial, as they clearly both have an impact on each other. Providers focusing on one specific aspect of psychological readiness may miss a key element in an athlete’s unique recovery process.

There are limitations to this study. First, this study only evaluated two time points following ACLR: 3.5 and 7 months. Changes in other time points, including recovery any time after 7 months post-ACLR, may offer additional information. In addition, adding a third time point would be useful in the future, allowing investigators to better determine the progression these values follow over time and their influence on each other. Secondly, the minimum sample size of 194 athletes for detecting weak effects was not met, but the observed effect size was moderate and detectable with the available sample. A larger sample size could have allowed for characteristic groupings or clusters of athletes that have similar traits. Additionally, using percent change rather than variance in raw scores could favor those who start with lower scores by giving them more room to improve. Finally, psychological and functional recovery are multifactorial processes influenced by external variables—such as rehabilitation quality, sport-specific exposure, or prior injury history—that were not controlled for in this study. These factors may have contributed variability to the findings and limit the generalizability of the results.

In conclusion, this study identified a statistically significant association between ACL-RSI and IKDC score changes from early to late recovery after ACLR. This parallel trend observed between the two assessments indicates that both psychological readiness and subjective knee function tend to progress in the same direction over ACL rehabilitation. These findings highlight the need to incorporate psychological preparation and support throughout all rehabilitation to promote both physical and mental recovery following an ACL injury.

## Data Availability

Data produced in the present study are available upon reasonable request to the authors.

## REFERENCES

1. Zink, G. P., Zwolski, C. M., Thomas, S. M., Paterno, M. V., & Schmitt, L. C. (2024). The Influence of Strength and Sport Exposure on Psychological Readiness After ACL Reconstruction. Sports health, 16(2), 239–246. DOI: 10.1177/19417381231223522

2. Webster, K. E., Nagelli, C. V., Hewett, T. E., & Feller, J. A. (2018). Factors Associated With Psychological Readiness to Return to Sport After Anterior Cruciate Ligament Reconstruction Surgery. The American journal of sports medicine, 46(7), 1545–1550. DOI: 10.1177/0363546518773757

3. Ardern, C. L., Österberg, A., Tagesson, S., Gauffin, H., Webster, K. E., & Kvist, J. (2014). The impact of psychological readiness to return to sport and recreational activities after anterior cruciate ligament reconstruction. British journal of sports medicine, 48(22), 1613–1619. DOI: 10.1136/bjsports-2014-093842

4. Lentz, T. A., Zeppieri Jr, G., George, S. Z., Tillman, S. M., Moser, M. W., Farmer, K. W., & Chmielewski, T. L. (2015). Comparison of physical impairment, functional, and psychosocial measures based on fear of reinjury/lack of confidence and return-to-sport status after ACL reconstruction. The American journal of sports medicine, 43(2), 345–353. DOI: 10.1177/0363546514559707

5. Zarzycki, R., Arhos, E., Failla, M., Capin, J., Smith, A. H., & Snyder-Mackler, L. (2021). Association of the Psychological Response to the ACL-SPORTS Training Program and Self-reported Function at 2 Years After Anterior Cruciate Ligament Reconstruction. The American journal of sports medicine, 49(13), 3495–3501. DOI: 10.1177/03635465211045388

6. Langford, J. L., Webster, K. E., & Feller, J. A. (2009). A prospective longitudinal study to assess psychological changes following anterior cruciate ligament reconstruction surgery. British journal of sports medicine, 43(5), 377–381. DOI: 10.1136/bjsm.2007.044818

7. Ashton, M. L., Kraeutler, M. J., Brown, S. M., & Mulcahey, M. K. (2020). Psychological Readiness to Return to Sport Following Anterior Cruciate Ligament Reconstruction. JBJS reviews, 8(3), e0110. DOI: 10.2106/JBJS.RVW.19.00110

8. Ohji, S., Aizawa, J., Hirohata, K., Ohmi, T., Mitomo, S., Koga, H., & Yagishita, K. (2022). Changes in subjective knee function and psychological status from preoperation to 6 months post anterior cruciate ligament reconstruction. Journal of experimental orthopaedics, 9(1), 114. DOI: 10.1186/s40634-022-00551-2

9. Suzuki, M., Ishida, T., Matsumoto, H., Kaneko, S., Inoue, C., Aoki, Y., Tohyama, H., & Samukawa, M. (2023). Association of Psychological Readiness to Return to Sports With Subjective Level of Return at 12 Months After ACL Reconstruction. Orthopaedic journal of sports medicine, 11(9), 23259671231195030. DOI: 10.1177/23259671231195030

10. Higgins, L. D., Taylor, M. K., Park, D., Ghodadra, N., Marchant, M., Pietrobon, R., Cook, C., & International Knee Documentation Committee (2007). Reliability and validity of the International Knee Documentation Committee (IKDC) Subjective Knee Form. Joint bone spine, 74(6), 594–599. DOI: 10.1016/j.jbspin.2007.01.036

11. Hopper, H. M., Bruce Leicht, A. S., Thompson, X. D., Gwathmey, F. W., Miller, M. D., Werner, B. C., Brockmeier, S. F., Diduch, D. R., & Hart, J. M. (2024). The effect of factors from different time points on psychological readiness following ACL reconstruction. Physical therapy in sport : official journal of the Association of Chartered Physiotherapists in Sports Medicine, 67, 161–166. DOI: 10.1016/j.ptsp.2024.05.002

12. Chmielewski, T. L., Jones, D., Day, T., Tillman, S. M., Lentz, T. A., & George, S. Z. (2008). The association of pain and fear of movement/reinjury with function during anterior cruciate ligament reconstruction rehabilitation. The Journal of orthopaedic and sports physical therapy, 38(12), 746–753. DOI: 10.2519/jospt.2008.2887

13. Truong, L. K., Mosewich, A. D., Holt, C. J., Le, C. Y., Miciak, M., & Whittaker, J. L. (2020). Psychological, social and contextual factors across recovery stages following a sport-related knee injury: a scoping review. British journal of sports medicine, 54(19), 1149–1156. DOI: 10.1136/bjsports-2019-101206

14. Kuenze, C., Bell, D. R., Grindstaff, T. L., Lisee, C. M., Birchmeier, T., Triplett, A., & Pietrosimone, B. (2021). A Comparison of Psychological Readiness and Patient-Reported Function Between Sexes After Anterior Cruciate Ligament Reconstruction. Journal of athletic training, 56(2), 164–169. DOI: 10.4085/1062-6050-0034.20

15. Baez, S., Harkey, M., Birchmeier, T., Triplett, A., Collins, K., & Kuenze, C. (2023). Psychological Readiness, Injury-Related Fear, and Persistent Knee Symptoms After Anterior Cruciate Ligament Reconstruction. Journal of athletic training, 58(11-12), 998–1003. DOI: 10.4085/1062-6050-0229.22

16. Milewski, M. D., Traver, J. L., Coene, R. P., Williams, K., Sugimoto, D., Kramer, D. E., Kocher, M. S., Micheli, L. J., Yen, Y. M., & Christino, M. A. (2023). Effect of Age and Sex on Psychological Readiness and Patient-Reported Outcomes 6 Months After Primary ACL Reconstruction. Orthopaedic journal of sports medicine, 11(6), 23259671231166012. DOI: 10.1177/23259671231166012

17. Kim, Y., Kubota, M., Sato, T., Inui, T., Ohno, R., & Ishijima, M. (2022). Psychological Patient-reported outcome measure after anterior cruciate ligament reconstruction: Evaluation of subcategory in ACL-Return to Sport after Injury (ACL-RSI) scale. Orthopaedics & Traumatology: Surgery & Research, 108(3), 103141. DOI: 10.1016/j.otsr.2021.103141

18. Webster, K. E., Feller, J. A., & Lambros, C. (2008). Development and preliminary validation of a scale to measure the psychological impact of returning to sport following anterior cruciate ligament reconstruction surgery. Physical therapy in sport : official journal of the Association of Chartered Physiotherapists in Sports Medicine, 9(1), 9–15. DOI: 10.1016/j.ptsp.2007.09.003

19. Higgins, L. D., Taylor, M. K., Park, D., Ghodadra, N., Marchant, M., Pietrobon, R., Cook, C., & International Knee Documentation Committee (2007). Reliability and validity of the International Knee Documentation Committee (IKDC) Subjective Knee Form. Joint bone spine, 74(6), 594–599. DOI: 10.1016/j.jbspin.2007.01.036

20. Daniel, A. V., Myer, G. D., Pashuck, T. D., & Smith, P. A. (2024). Low Preoperative Brief Resilience Scale Scores Are Associated With Inferior Preoperative and Short-Term Postoperative Patient Outcomes Following Primary Autograft Anterior Cruciate Ligament Reconstruction. Arthroscopy, Sports Medicine, and Rehabilitation, 6(1), Article 100858. DOI: 10.1016/j.asmr.2023.100858

21. Barth, T., Bond, C. W., MacFadden, L. N., Skelley, N. W., Combs, J., & Noonan, B. C. (2024). Effect of Time and Sex on Post-Anterior Cruciate Ligament Reconstruction Psychological Patient-Reported Outcome Measure Scores. Journal of athletic training, 59(9), 898–905. DOI: 10.4085/1062-6050-0189.23

